# Temporal changes in sleep parameters and body mass index after using the Pokémon Sleep app

**DOI:** 10.1101/2024.06.29.24309336

**Authors:** Masao Iwagami, Jaehoon Seol, Masashi Yanagisawa

## Abstract

Pokémon Sleep is a new sleep-tracking game app released in July 2023. We examined whether sleep parameters would improve with the use of the Pokémon Sleep app, and whether participants’ improvement status was associated with changes in their body mass index (BMI). We analyzed data from 2,063 volunteer Japanese participants (mean age 38.3 ± 10.7 years, female 82.1%) who used both the Pokémon Sleep and Asken (a commonly-used app in Japan to record daily BMI) for ≥90 days. During the 90 days, improvement was seen in total sleep time (TST) for 44.8% of participants, sleep latency for 18.1%, and percentage of wakefulness after sleep onset for 24.4%. BMI tended to decrease faster in participants with improved TST and sleep latency. In conclusion, the Pokémon Sleep app can improve some sleep parameters, and by association, lead to improvement in BMI.

## Introduction

In the era of digital health, a variety of mobile apps have been developed to record daily health-related parameters (e.g., step counts and screen time), with some including game elements to nudge users towards better health behaviors. Among these apps are a small number designed to track sleep behaviors and characteristics; however, their influence on users’ behavioral and clinical outcomes remains unknown [1, 2].

Pokémon Sleep (The Pokémon Company, Tokyo, Japan) is a sleep-tracking app released in July 2023 in multiple countries, including Japan and the United States. Compared to previous sleep-tracking apps [1, 2], Pokémon Sleep has a stronger game element, rewarding users with points for longer sleep and the ability to raise Pokémon. Therefore, it can be hypothesized that key sleep parameters, such as total sleep time (TST), sleep latency (the duration from the start time in bed to sleep onset), and wakefulness after sleep onset (WASO), will improve after starting Pokémon Sleep. Further, we hypothesized that people whose sleep improved after starting Pokémon Sleep would also have improved health indicators, specifically, body mass index (BMI).

To examine these hypotheses, we conducted a retrospective observational study of data from users of both Pokémon Sleep and Asken (asken, Inc., Tokyo, Japan), an app commonly used in Japan to record daily diets and BMI [3].

## Methods

Details of the Pokémon Sleep and Asken apps are described elsewhere [3, 4]. In brief, Pokémon Sleep tracks and records daily sleep characteristics using the accelerometer functions of mobile phones or smart devices named Pokémon GO Plus+. Users are instructed to keep the app open and place the device by their pillow, or other location where it will detect only the user’s body movements. Using algorithms established by Cole and Kripke [5], Pokémon Sleep distinguishes between periods of sleep and wakefulness. From these records, we calculated daily TST, sleep latency, and the proportion of WASO (%WASO) during TST. Asken is a calorie and nutrition tracker that also monitors daily BMI.

In-app messages were sent to all Asken users from January 19 through 31, 2024, asking those who used both Pokémon Sleep and Asken for permission to use their stored data. Respondents were asked to provide their Pokémon Sleep user IDs in their electronic informed consent so that the Pokémon Sleep and Asken data were linkable. The study was approved by the Institutional Review Board of Sapporo Yurinokai Hospital, Japan.

Using the retrieved data, we identified when each participant had started using Pokémon Sleep (“day 0”), and included those continuing it until day 89, with or without gaps. Persons were excluded if they had not used Asken during the 90 days of Pokémon Sleep recordings, or did not have BMI recordings at days 0 and 89 (i.e., the first and last dates). For gaps or days when the participants did not use Pokémon Sleep or Asken, missing data were imputed linearly from the records immediately before and after the day.

In statistical analysis, we summarized the basic characteristics of participants including age, sex, and baseline BMI. We also cross-sectionally estimated cubic spline curves for each sleep parameter (days 0–6 average) with BMI on day 0, based on a generalized additive model. Second, we plotted the average trend of each sleep parameter from days 0 to 89. For each sleep characteristic, we then classified participants into two groups, improved/not improved, based on the slope calculated for individuals during the 90 days. We regarded a slope >0 as an improvement in TST, and a slope <0 as an improvement in sleep latency and %WASO. Finally, for each sleep parameter, we plotted the average change in BMI from days 0 to 89 by group. Repeated-measures analysis of variance (ANOVA) at days 0, 29, 59, and 89 was used to test whether the temporal trend in BMI differed between the groups, adjusting for age and sex.

All analyses were performed using Python (version 3.12.2), which was built into Visual Studio Code (version 1.87.2). Statistical significance was set at P<0.05 (2-tailed).

## Results

Of 6,052 responses to our request, 3,934 participants used Pokémon Sleep for ≥90 days, and 2,063 participants were included for the analysis (**Supplementary Figure 1**). Of the 90 days, the mean number of days spent on Pokémon Sleep and Asken was 64.1 (standard deviation [SD] 16.6 days) and 59.6 (SD 28.6 days), respectively. Participants’ baseline characteristics are shown in **Supplementary Table 1**. In the cross-sectional analysis (**Supplementary Figure 2**), longer TST (up to around 8.5 hours) and shorter sleep latency were associated with lower BMI.

On average, the TST increased from days 0 to 89 by approximately 0.5 hours, whereas sleep latency and %WASO were nearly constant (**Supplementary Figure 3**). However, calculating the slope for individuals, 924 (44.8%), 374 (18.1%), and 504 (24.4 %) participants were classified as “improved” for TST, sleep latency, and %WASO, respectively. Participants’ baseline characteristics by improvement status are shown in **Supplementary Table 2**.

**Figure 1** shows the temporal changes in BMI by improvement status groups. Regardless of the groups, participants’ BMI tended to decrease during the 90 days. However, participants with improved TST (**panel A**) showed a steeper and more sustained decline in BMI than did the non-improved group, although the difference was not statistically significant in the repeated-measures ANOVA (P=0.107). The mean change in BMI at day 89 was -0.38 and -0.19, respectively (unadjusted difference=-0.19, 95% confidence interval [CI]: -0.40–0.03; age-sex-adjusted difference=-0.25, 95% CI: -0.51–0.01). Participants with improved sleep latency (**panel B**) showed a significantly faster decline in BMI than the non-improved group (P=0.030). The mean change in BMI at day 89 was -0.51 and -0.23, respectively (unadjusted difference=-0.28, 95% CI: -0.56–0.00; age-sex-adjusted difference=-0.36, 95% CI: -0.69–-0.02). There was no significant difference (P=0.851) in the amount of BMI change by %WASO improvement status (**panel C**).

**Figure 1.**
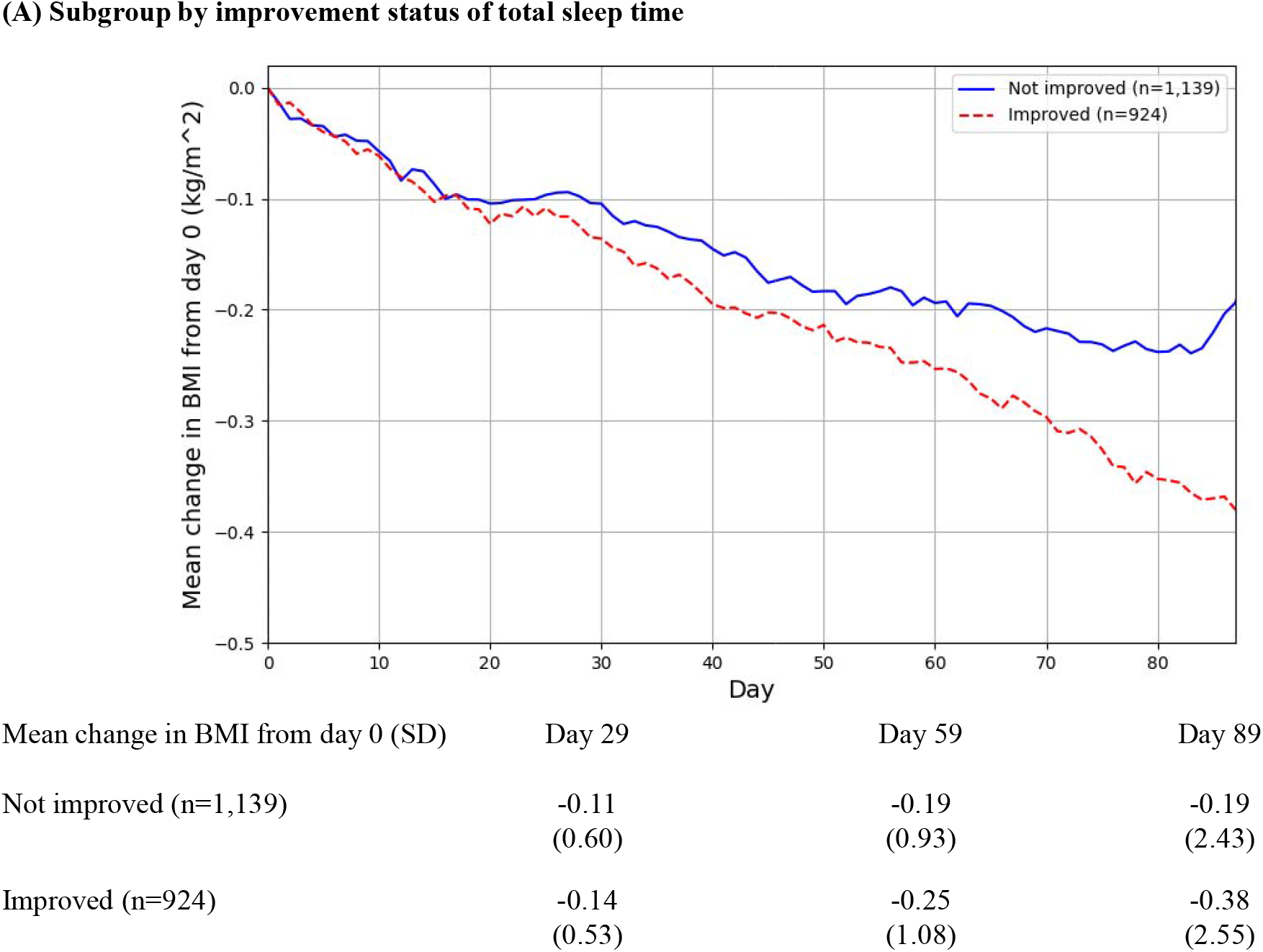
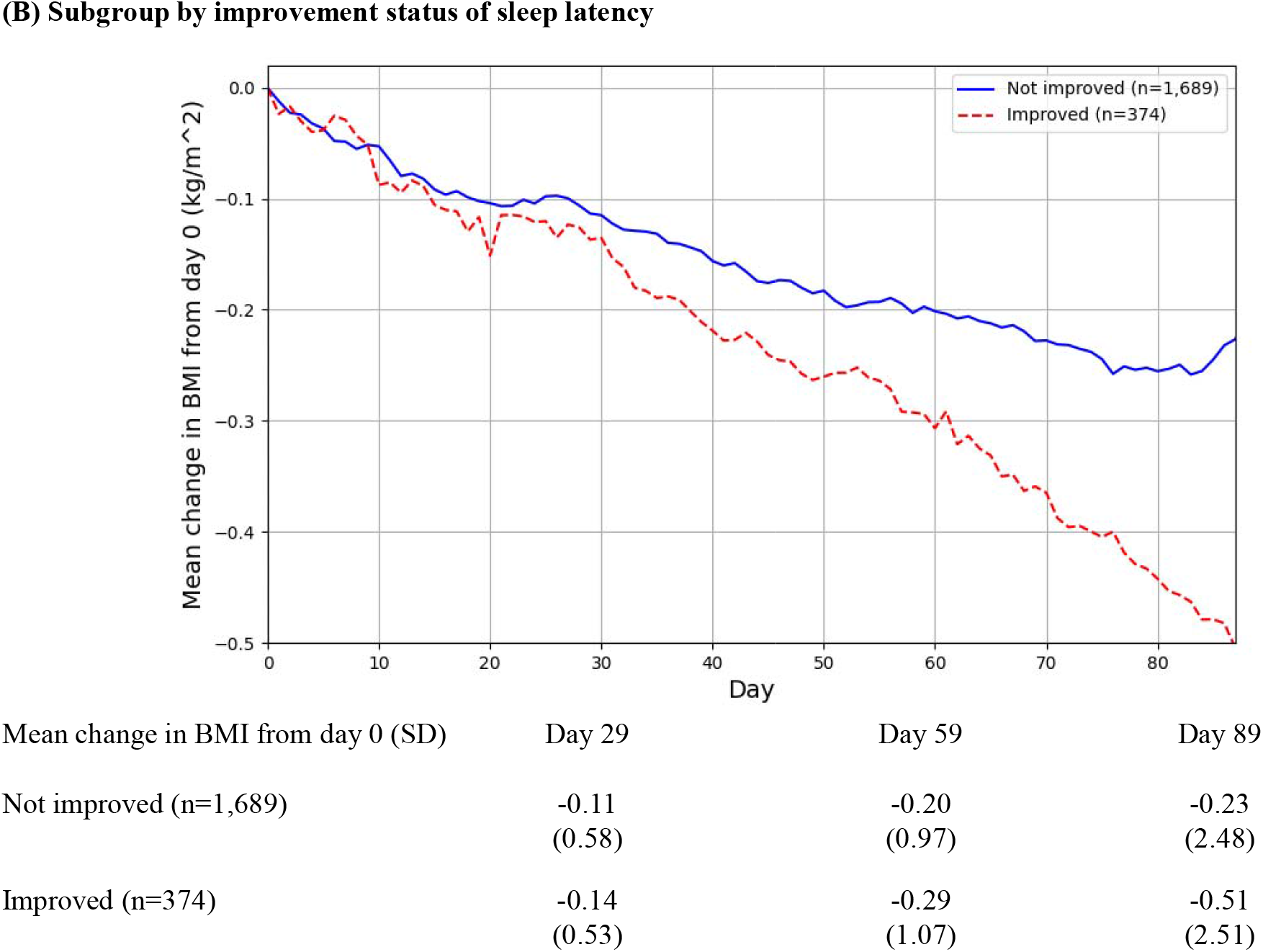
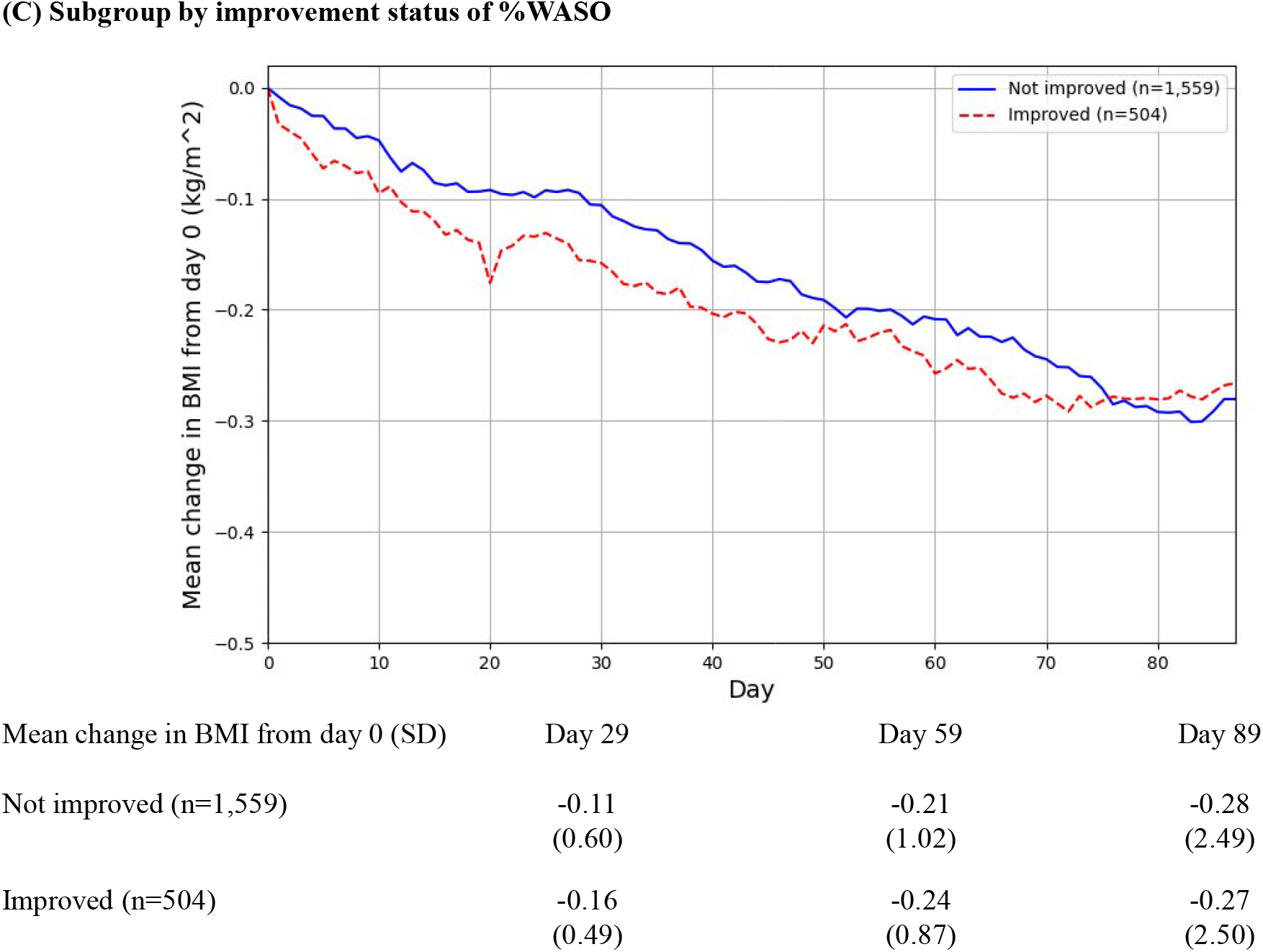
Temporal changes in body mass index by improvement status of total sleep time (panel A), sleep latency (panel B), and % wakefulness after sleep onset (panel C) BMI, body mass index; %WASO, percentage of wakefulness after sleep onset; SD, standard deviation.

## Discussion

In this study of 2,063 volunteer participants using both Pokémon Sleep and Asken for ≥90 days, improvement was seen in TST for 44.8% of participants, sleep latency for 18.1%, and %WASO for 24.4%. Participants’ BMI tended to decrease during the 90 days, most likely because Asken users are typically motivated to control their diets and decrease their body weight. However, BMI tended to decrease faster in participants with improved TST (without a statistical significance) and sleep latency (with a statistical significance). Potential mechanisms may include improvement of appetite-related hormones (e.g., ghrelin and leptin), or changes in energy metabolism and balance [6, 7].

This study had several limitations. The Pokémon Sleep app was the first to use the accelerometer functions of mobile phones to measure sleep on the bed. To our knowledge, its validity in distinguishing between sleep and wakefulness has not been rigorously tested in comparison to polysomnography or actigraphy. Second, those who used both Pokémon Sleep and Asken for ≥90 days may be highly health-conscious and the generalizability of our findings to more general populations is thus unknown.

Despite these limitations, to our knowledge, this is the first evidence in the general population that a sleep-tracking app can improve some sleep parameters, and by association, lead to improvement in physical health parameters (i.e., BMI). Further studies are warranted to further examine whether the use of sleep-tracking apps can improve sleep and physical health parameters, and ultimately, their long-term health.

## Acknowledgements

We express our gratitude to the personnel of The Pokémon Company (Tokyo, Japan), asken Inc. (Tokyo, Japan), and S’UIMIN Inc. (Tokyo, Japan) for their contributions to data preparation. We also thank Editage (www.editage.com) for the English language editing.

## Data Availability Statement

The data underlying this article cannot be shared publicly for the privacy of individuals that participated in the study. The data may be shared on reasonable request to the corresponding author.

## Supplemental files

**Supplementary Figure 1. Participant flow chart**

**Supplementary Table 1. Baseline characteristics of study participants**

**Supplementary Figure 2. Cross-sectional associations between sleep parameters and body mass index at baseline**

**Supplementary Figure 3. Temporal changes in sleep parameters, overall and by improvement status**

**Supplementary Table 2. Baseline characteristics of study participants by improvement status for sleep parameters**

