## Supplementary Tables 1, 2, and Figures 1-3 for "Temporal changes in sleep parameters and body mass index after using the Pokémon Sleep app"

**Supplementary Figure 1. Participant flow chart**

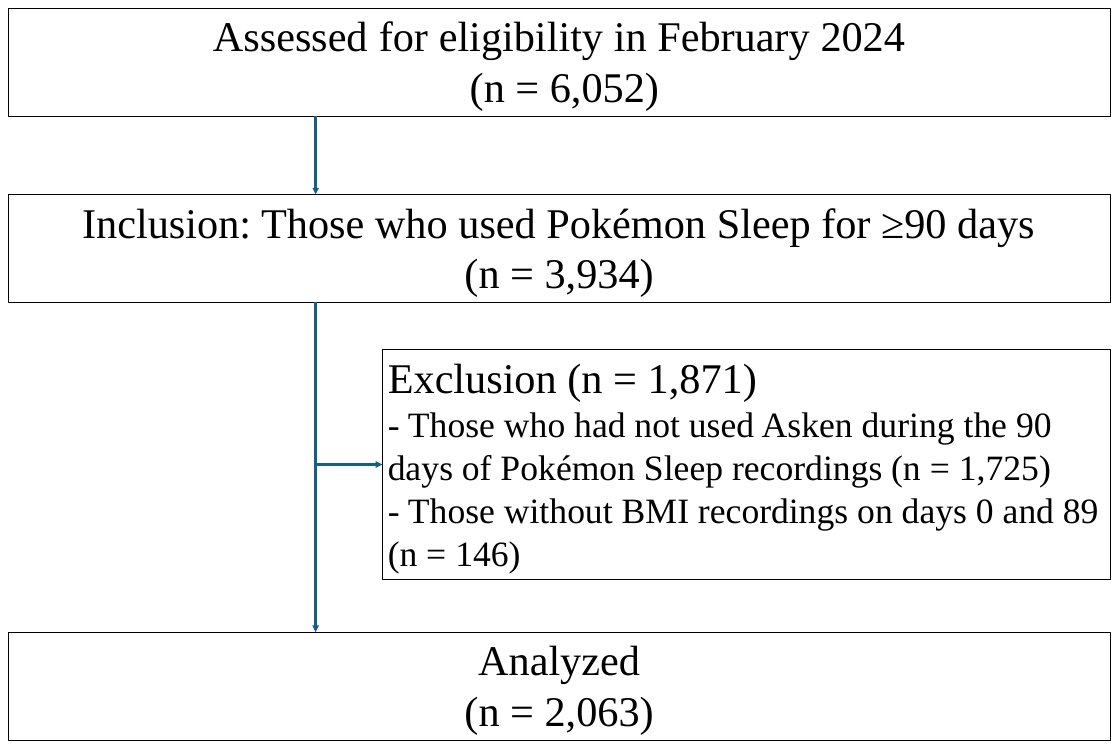

**Supplementary Table 1. Baseline characteristics of study participants**

| Variables | Study participants (N＝2,063) |
| --- | --- |
| Age (years) | 38.3 ± 10.7 |
| No. of female (%) | 1,694 (82.1) |
| BMI (day 0), kg/m^2^ | 24.1 ± 4.4 |
| Total sleep time*, hour | 6.2 ± 1.2 |
| Sleep onset latency*, min | 19.2 ± 14.2 |
| %WASO*, % | 9.1 ± 8.2 |
| BMI, body mass index; %WASO, percentage of wakefulness after sleep onset.  *Days 0–6 average. | |

**Supplementary Figure 2. Cross-sectional associations between sleep characteristics and body mass index at baseline**

(A) **Association between total sleep time (days 0-6 average) and BMI (day 0)**

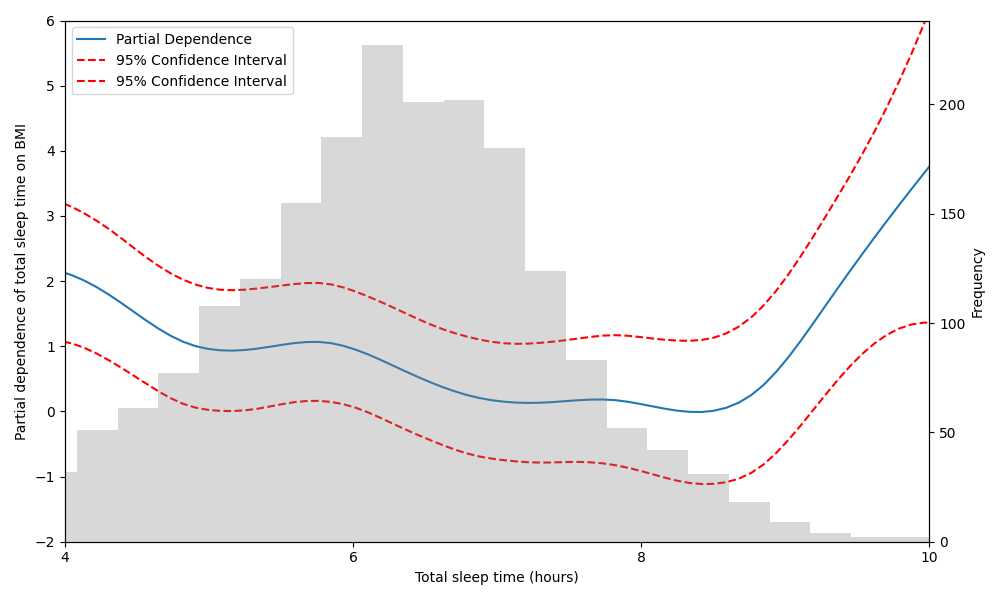

(B)
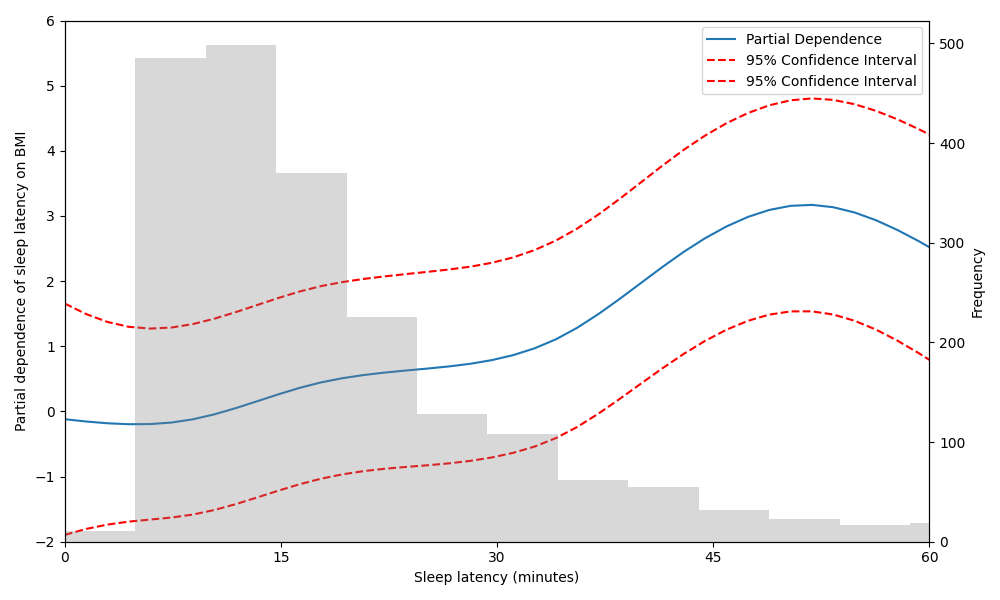
**Association between sleep latency (days 0-6 average) and BMI (day 0)**
(C) **Association between %WASO (days 0-6 average) and BMI (day 0)**

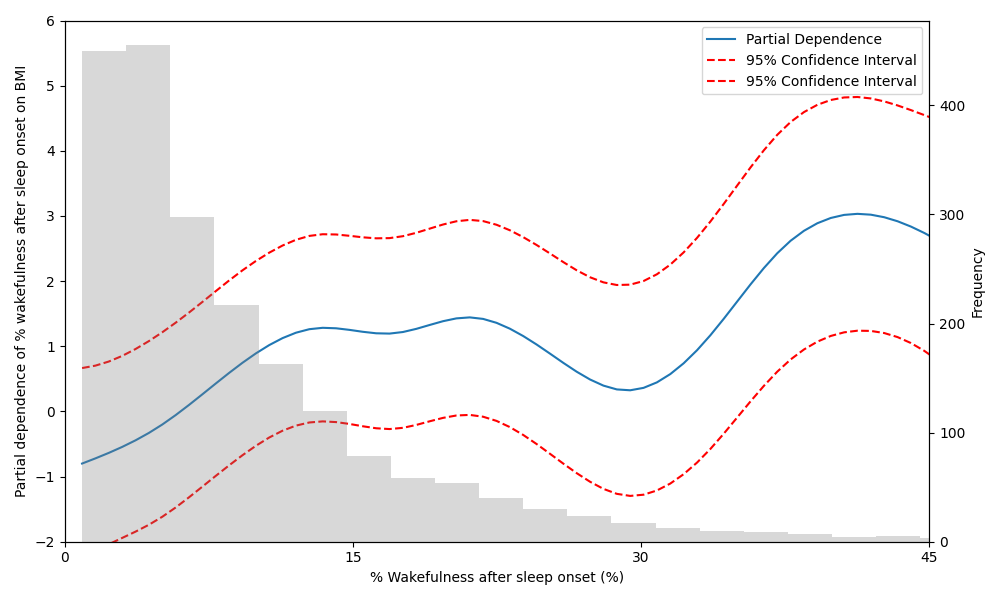

**Supplementary Figure 3. Temporal changes in sleep parameters, overall and by improvement status**

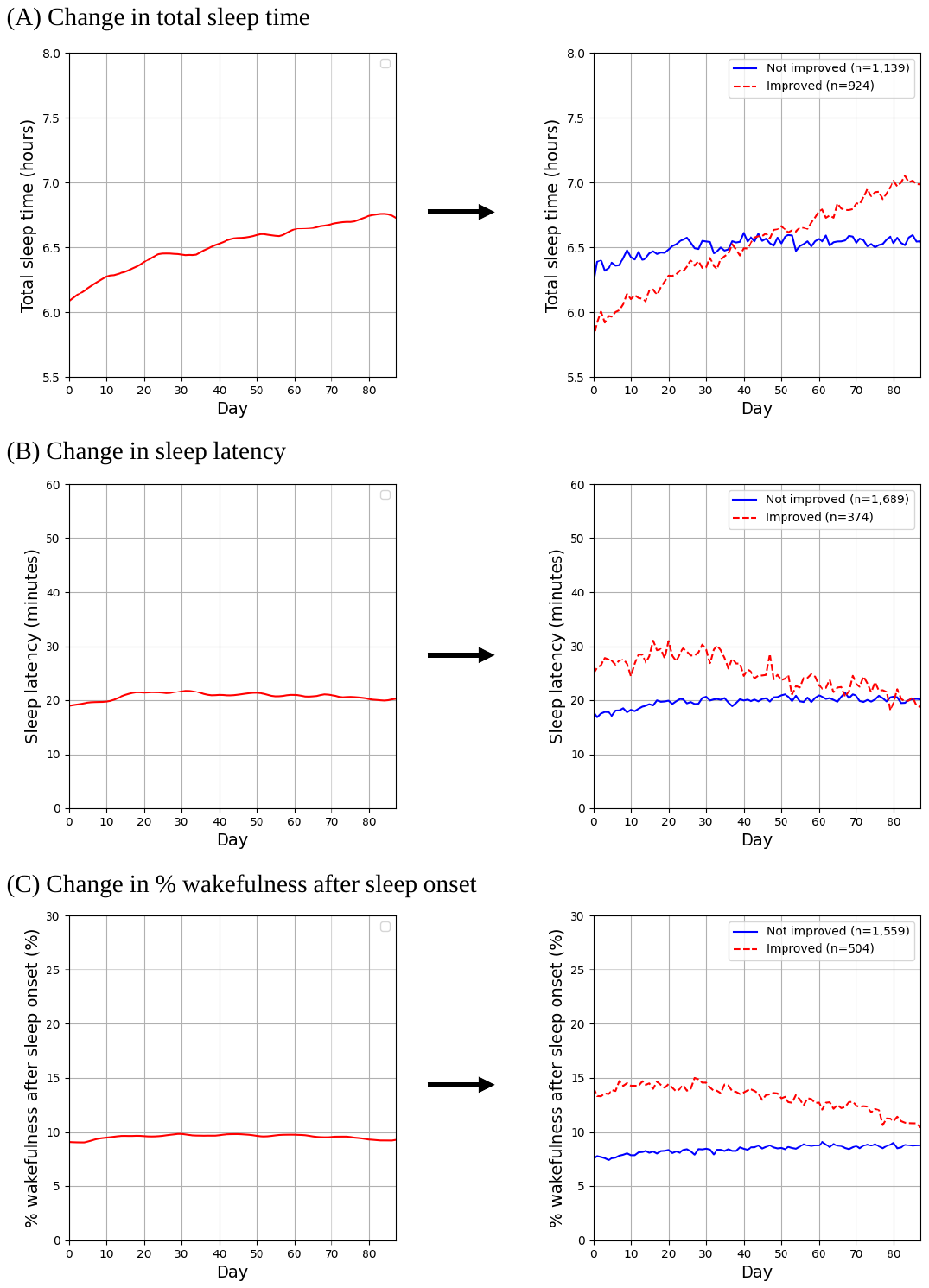

**Supplementary Table 2. Baseline characteristics of study participants by improvement status for sleep parameters**

(A) **Subgroup by improvement status of total sleep time**

| Variables | Improved (n=924) | Not improved (n=1,139) |
| --- | --- | --- |
| Age (years) | 38.5 ± 10.6 | 38.1 ± 10.7 |
| No. of female (%) | 730 (79.0) | 964 (84.6) |
| BMI (day 0), kg/m^2^ | 24.3 ± 4.5 | 24.0 ± 4.4 |
| Total sleep time*, hour | 5.9 ± 1.3 | 6.4 ± 1.2 |
| Sleep onset latency*, min | 20.8 ± 15.2 | 17.9 ± 13.2 |
| %WASO*, % | 9.9 ± 8.7 | 8.4 ± 7.7 |
| BMI, body mass index; %WASO, percentage of wakefulness after sleep onset.  *Days 0–6 average. | | |

(B) **Subgroup by improvement status of sleep onset latency**

| Variables | Improved (n=374) | Not improved (n=1,689) |
| --- | --- | --- |
| Age (years) | 38.4 ± 10.9 | 38.2 ± 10.6 |
| No. of female (%) | 288 (77.0) | 1406 (83.2) |
| BMI (day 0), kg/m^2^ | 24.1 ± 4.3 | 24.1 ± 4.5 |
| Total sleep time*, hour | 6.2 ± 1.2 | 6.2 ± 1.2 |
| Sleep onset latency*, min | 26.8 ± 18.3 | 17.5 ± 12.5 |
| %WASO*, % | 11.5 ± 9.2 | 8.6 ± 7.8 |
| BMI, body mass index; %WASO, percentage of wakefulness after sleep onset.  *Days 0–6 average. | | |

(C) **Subgroup by improvement status of % wakefulness after sleep onset**

| Variables | Improved (n=504) | Not improved (n=1,559) |
| --- | --- | --- |
| Age (years) | 38.8 ± 11.0 | 38.1 ± 10.5 |
| No. of female (%) | 408 (81.0) | 1,286 (82.5) |
| BMI (day 0), kg/m^2^ | 24.3 ± 4.5 | 24.1 ± 4.4 |
| Total sleep time*, hour | 6.2 ± 1.2 | 6.2 ± 1.3 |
| Sleep onset latency*, min | 22.4 ± 15.1 | 18.2 ± 13.7 |
| %WASO*, % | 13.7 ± 9.7 | 7.6 ± 7.0 |
| BMI, body mass index; %WASO, percentage of wakefulness after sleep onset.  *Days 0–6 average. | | |
